# Examining Clinical Reasoning during Stimulability Testing for Voice-Specialized Speech-Language Pathologists: A Qualitative Study

**DOI:** 10.64898/2025.12.08.25341853

**Authors:** Elizabeth D. Young

**Affiliations:** School of Behavioral and Brain Sciences, University of Texas at Dallas, Richardson, TX

## Abstract

**Purpose:** Stimulability testing is a wide-spread and highly valued behavioral assessment tool for voice-specialized speech-language pathologists (SLPs). However, there is currently no research examining how voice SLPs use stimulability testing to inform their clinical reasoning process and decisions. The purpose of this qualitative study was to examine the clinical reasoning underlying stimulability testing for voice-specialized SLPs across the experience spectrum.

**Methods:** Semi-structured interviews were conducted with eight voice-specialized SLPs (four early-career, four late-career) regarding stimulability testing, including how they used stimulability testing to form clinical conclusions such as candidacy and prognosis for behavioral therapy. Interviews were transcribed and analyzed using the interpretative phenomenological analysis (IPA) framework (Smith & Osborn, 2003).

**Results:** Five themes emerged from the IPA analyses: *Perceiving and monitoring patient responses; Developing and trusting clinical skills; Decision-making strategies; Drawing clinical conclusions;* and *Fostering a purposeful therapeutic relationship.* Within the *Drawing clinical conclusions* theme, clinicians differed on the utility of stimulability testing as a tool for determining patient candidacy and prognosis for behavioral therapy.

**Conclusion:** SLPs rely on both analytical and intuitive methods of clinical reasoning during stimulability testing. However, the lack of research tying stimulability testing to clinical outcomes has led to an overreliance on intuitive reasoning when SLPs attempt to draw clinical conclusions. Further empiric support for the clinical functions of stimulability testing is needed to support the clinical reasoning process surrounding this assessment tool.

## Introduction

Stimulability testing is the assessment of an individual’s ability to change or modify a behavior, typically towards “normal”, utilizing therapeutic techniques or facilitators. Recent survey-based research has found that the majority of voice-specialized speech-language pathologists (SLPs) and laryngologists conduct stimulability testing routinely with their voice patients (Coleman et al., 2025; Toles & Young, 2023). Additionally, 97% of these same SLPs considered stimulability testing important to their practice, and 100% of laryngologists reported a benefit from performing stimulability testing. SLPs and laryngologists report performing stimulability testing to inform a variety of clinical decisions, ranging from diagnostic (e.g., differential diagnosis of muscle tension dysphonia versus spasmodic dysphonia), to prognostic (e.g., estimating a patient’s prognosis for improvement with behavioral intervention), to therapeutic (e.g., evaluating and potentially improving the patient’s motivation for behavioral therapy; Coleman et al., 2025; Toles et al., 2024; Toles & Young, 2023). Stimulability *testing* (i.e., the assessment procedure) is also used to make determinations regarding the patient’s stimulability *level* (i.e., the results of the procedure – the degree to which the patient is “stimulable”). Taken together, these factors indicate that stimulability testing is a ubiquitous and multifunctional clinical tool for the assessment of voice disorders.

Despite the importance and pervasiveness of stimulability testing, there is very limited standardization of stimulability testing practices. Literature describing stimulability testing procedures is generally descriptive in nature, and most practicing SLPs learn to conduct stimulability testing via a mentorship model (Toles & Young, 2023). As a result, stimulability testing conducted by one clinician may differ substantially from stimulability testing conducted by another clinician. As noted above, however, stimulability testing is considered by both SLPs and laryngologists to be an important element to a behavioral voice evaluation that can impact both diagnostic and treatment decisions. Thus, there is a great need for research into not only the stimulability testing process itself, but also the processes underlying clinical reasoning that occurs during stimulability testing that allows voice care specialists to make treatment decisions.

The current study utilized semi-structured interviews to examine the clinical reasoning process and decisions made during stimulability testing for eight voice-specialized SLPs across the clinical experience spectrum. Literature examining clinical reasoning for speech-language pathologists is extremely limited, particularly in speciality areas such as the treatment of voice disorders. Given both the ubiquity and heterogeneity of stimulability testing within the behavioral voice evaluation, however, a greater understanding of the reasoning process used in stimulability testing is warranted. Thus, the current study had the following research questions:

1. How do voice-specialized speech-language pathologists across the experience spectrum describe their clinical reasoning process surrounding stimulability testing?
2. How does the clinical reasoning process described by voice-specialized speech-language pathologists during stimulability testing inform clinical decisions such as patient stimulability level, diagnosis, and candidacy and prognosis for behavioral therapy?

Gaining a deeper understanding of the reasoning process that occurs during stimulability testing and the subsequent decisions informed by these processes will lay the groundwork for creating a more standardized framework for stimulability testing, which in turn could be used to improve patient care outcomes, increase the predictive validity of stimulability testing, and improve clinical education for new SLPs.

### Clinical Reasoning

The term “clinical reasoning” is used throughout this paper to refer to the process and thinking patterns used to integrate and evaluate clinical data (e.g., patient history, patient presentation, response to stimulability testing, etc.) to make both diagnostic and treatment decisions. This definition aligns closely with a recent definition posed in the Journal of Professional Nursing developed based on expert consensus: “Clinical decision making is a contextual, continuous, and evolving process, where data are gathered, interpreted, and evaluated in order to select an evidence-based choice of action.” (Tiffen et al., 2014, p. 401). Clinical reasoning is not a one-time moment but a continually evolving process of evaluating, re-evaluating, and integrating new information to deliver the best possible care to the patient.

Clinical reasoning has been given many different names and has been examined through multiple lenses in the literature. A 2020 scoping review of clinical reasoning across health professions found 110 different terms used to describe clinical reasoning (M.E. Young et al., 2020). This same review grouped terminology into six overarching categories: reasoning skills, reasoning performance, reasoning process, the outcome of reasoning, the context of reasoning, and the purpose/goal of reasoning. The literature regarding clinical reasoning in speech-language pathology is relatively sparse. However, the existing literature generally falls into the outcome of reasoning, reasoning performance, and reasoning process categories. While literature in all these areas will be reviewed, it should be noted that the focus of the current study lies primarily in the process of reasoning, or the “component processes by which reasoning unfolds” (M.E. Young et al., 2020, p. 6).

### Clinical Reasoning Research in Speech-Language Pathology

The above cited scoping review found that across 625 papers spanning 18 different health professions, only a single article was specific to speech-language pathology (M. E. Young et al., 2020). Authors have called for more literature on this topic, especially given the potential impact clinical reasoning research could have on the education of new SLPs (McAllister & Rose, 2000). Despite this call, research on clinical reasoning specific to speech-language pathology remains limited. Of note, no literature specific to the assessment of voice disorders or stimulability assessment was found in the current literature search, highlighting the need for increased literature regarding clinical reasoning with this population and within the field of speech-language pathology as a whole.

Clinical reasoning skills have been shown to develop throughout both formal training and with clinical experience in speech-language pathology. Studies examining clinical reasoning skills in students found that they often have difficulty choosing appropriate assessments, organizing and interpreting assessment information, planning diagnostic strategies, and conceptualizing problems at an abstract level (Hoben et al., 2007; Howarth et al., 2005). While graduate students improve in diagnostic accuracy and appropriate recommendations for therapy throughout graduate education, they did not improve in their ability to formulate an accurate hypothesis and select appropriate evaluation instruments (Dudding & Pfeiffer, 2018). Compared to novice clinicians, experienced clinicians are more likely have specific assessment and treatment plans and draw more heavily on previous clinical experience when presented with case studies (Ginsberg et al., 2016; Sauerwein & Wegner, 2020). These studies support the idea that experience is necessary for gaining the ability to see and respond to patterns in patient presentation.

Clinical reasoning outcomes (i.e., diagnostic accuracy) have been examined for various communication disorders, including speech sound disorders (Diepeveen et al., 2020) and language impairments (Records & Tomblin, 1994; Selin et al., 2019). Results show that while speech-language pathologists are generally reliable with one another in their broad diagnosis categories, discrepancies exist in the language used to describe a diagnostic category.

Furthermore, several studies showed that outside influences (e.g., resource availability, local regulations, etc.) often profoundly influenced both diagnostic and treatment decisions made by speech-language pathologists (Diepeveen et al., 2020; Selin et al., 2019). For instance, SLPs surveyed by Diepeveen et al. (2020) overwhelmingly reported that the availability of or familiarity with a certain diagnostic test was a primary driver in deciding which tests to use. No literature exists to the author’s knowledge examining the relationship between clinical reasoning and diagnostic outcomes for other diagnostic categories, including voice disorders.

Extremely limited literature exists examining the clinical reasoning process specific to speech-language pathologists. Indeed, much of the literature that does exist uses existing terminology and frameworks from other healthcare professions, with examples and applications to SLP-specific disorders (Campbell, 1998; McAllister & Rose, 2000). While it is true that the general process is likely very similar between healthcare professions, there are doubtless subtleties of clinical reasoning that are specific to SLPs, particularly among specialized areas of treatment such as voice disorders. Stimulability testing, in particular, is a relatively unique technique to speech-language pathology. There is therefore a need to complete additional clinical reasoning research in specialized areas of speech-language pathology to determine if and where differences exist that are specific to the field.

### Frameworks for Clinical Reasoning

Many theories or frameworks describing the process and development of clinical reasoning have emerged in the literatures of various healthcare professions such as medicine, nursing, and physical therapy. These frameworks can generally be sorted into analytical, intuitive, or hybrid/combination frameworks. Analytical frameworks of clinical reasoning describe the conscious, deliberate reasoning process utilized by healthcare professionals when making diagnostic or treatment decisions. Intuitive frameworks, in contrast, emphasize intuitive decision-making based on the fast, often subconscious information-gathering that occurs during a patient-provider interaction. Combination frameworks rely on both types of processes, which may be weighed differentially depending on the context. For instance, Croskerry (2009) proposed a framework that suggests that the closer a patient’s fit is to the clinician’s mental model of a particular disorder, the stronger intuitive reasoning will be – but when a patient’s presentation is more ambiguous, analytical reasoning takes over as the primary reasoning process. A single hybrid framework known as the “contextualized clinical reasoning” framework emerged recently as a framework for speech-language pathologists (Pillay & Pillay, 2021). This framework suggests that clinical reasoning is a relationship between patient assessment results, context (e.g., patient’s socioeconomic status, resource availability, etc.), healthcare practitioner characteristics, and auxiliary resources such as peer support and multidisciplinary collaboration. This framework therefore acknowledges the impact of both patient and clinician factors, centering it within the overarching evidence-based practice framework utilized by the American Speech-Language Hearing Association (described further below).

## Methods

### Study Design and Methodology

Qualitative methods were chosen for this study, as they are uniquely suited to exploring individual experiences and examining thought processes. Further, most of the studies examining clinical reasoning in speech-language pathology have used qualitative methods (Dudding & Pfeiffer, 2018; Ginsberg et al., 2016; Hoben et al., 2007; Howarth et al., 2005; Sauerwein & Wegner, 2020). The American Speech-Language-Hearing Association (ASHA) Evidence-Based Practice (EBP; Straus et al., 2018) framework was selected as the overarching conceptual framework for this study, as the sampling methods of the study required all participants to be ASHA-certified (or in the process of receiving certification) and therefore familiar with this framework. This framework recommends the integration of clinical expertise/expert opinion, internal and external evidence, and client, patient, and caregiver perspectives for making clinical decisions (ASHA, n.d.). The author engaged in memoing and frequent reflexivity throughout the study process to increase transparency and rigor; her positionality statement generated both during the project and after project completion is available for review in the supplemental materials.

The interpretative phenomenological analysis (IPA) was selected as the guiding methodology for the study (Larkin et al., 2021; Smith & Osborn, 2003). IPA has its foundations in three areas of psychology; hermeneutics (the theory and methodology of interpretation), ideography (the study of how individuals make sense of situations), and phenomenology (the study of clarifying and elucidating a phenomenon). From an axiological standpoint, IPA places emphasis on value of those who have personal experience with the situation being studied (in this case, practicing clinicians who have experience performing stimulability testing for voice disorders). During data analysis, IPA emphasizes the use of a double-hermeneutic approach, where there is a focus on trying to make sense of both the participant’s experience and the researcher’s interpretation of their experience. Thus, from an ontological and epistemological approach, knowledge and reality are both formed at the individual, personal level, but also subject to interpretation between individuals. IPA is a useful paradigm for use when trying to understand an individual’s experience, how they make meaning from this experience, as well as when introducing outside interpretations of these individual’s experiences. The methods outlined in the current published IPA guidelines (e.g., Larkin et al., 2021) were used to guide the analysis of all semi-structured interviews collected in the study.

### Sampling Methods

Eight voice-specialized speech-language pathologists (referred to hereafter as “clinicians”) were recruited as participants for the study. Four clinicians were recruited as “early-career”, which was defined as having less than four years of full-time practice experience including their clinical fellowship, and four were recruited as “late-career”. These clinicians must have recently (i.e., within the last five years) provided clinical care to patients with voice disorders, have received at least 3 months of specialized training in the assessment and treatment of voice disorders, hold or be eligible to hold the ASHA certificate of clinical competence for speech-language pathology, and be currently licensed to practice as a speech-language pathologist. Clinicians were purposefully sampled across a broad range of experience levels and training backgrounds (see Table 1) so a variety of points of view regarding stimulability testing would be represented.

**Table 1.**
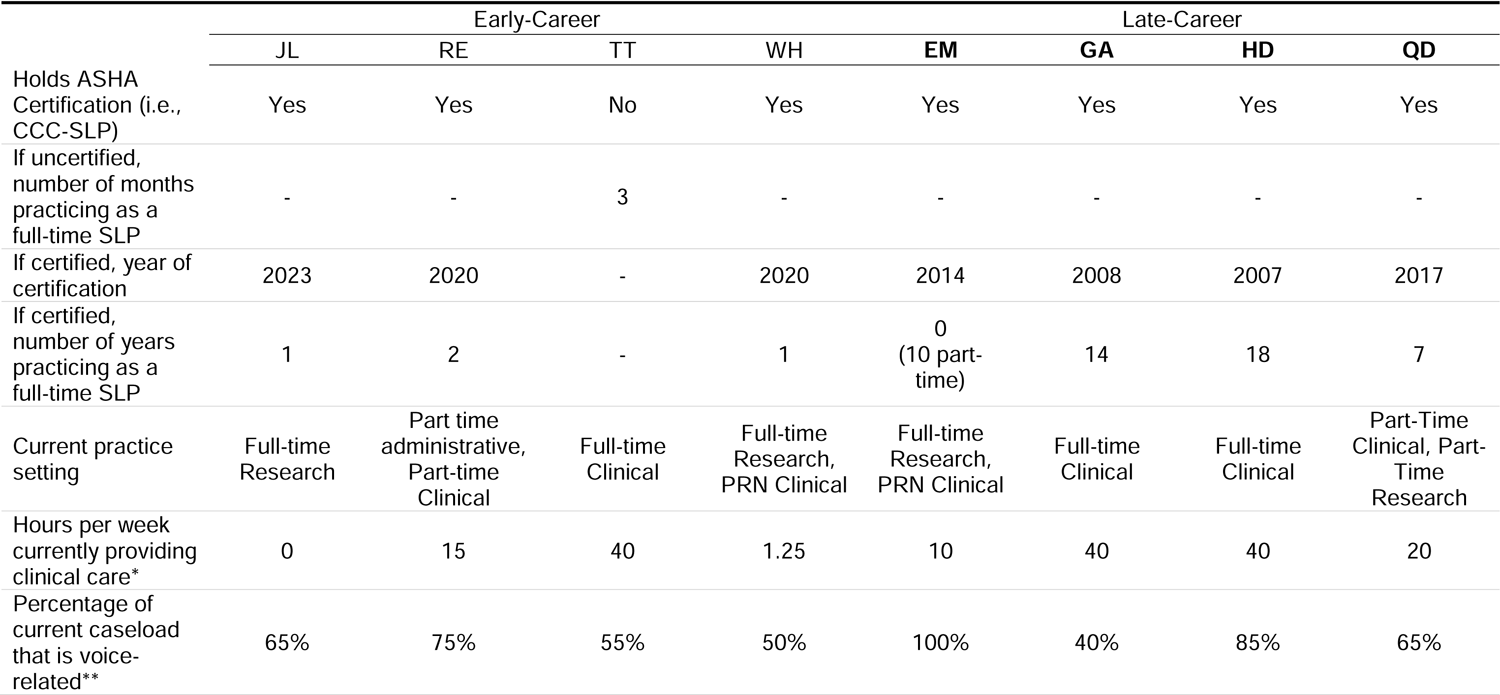
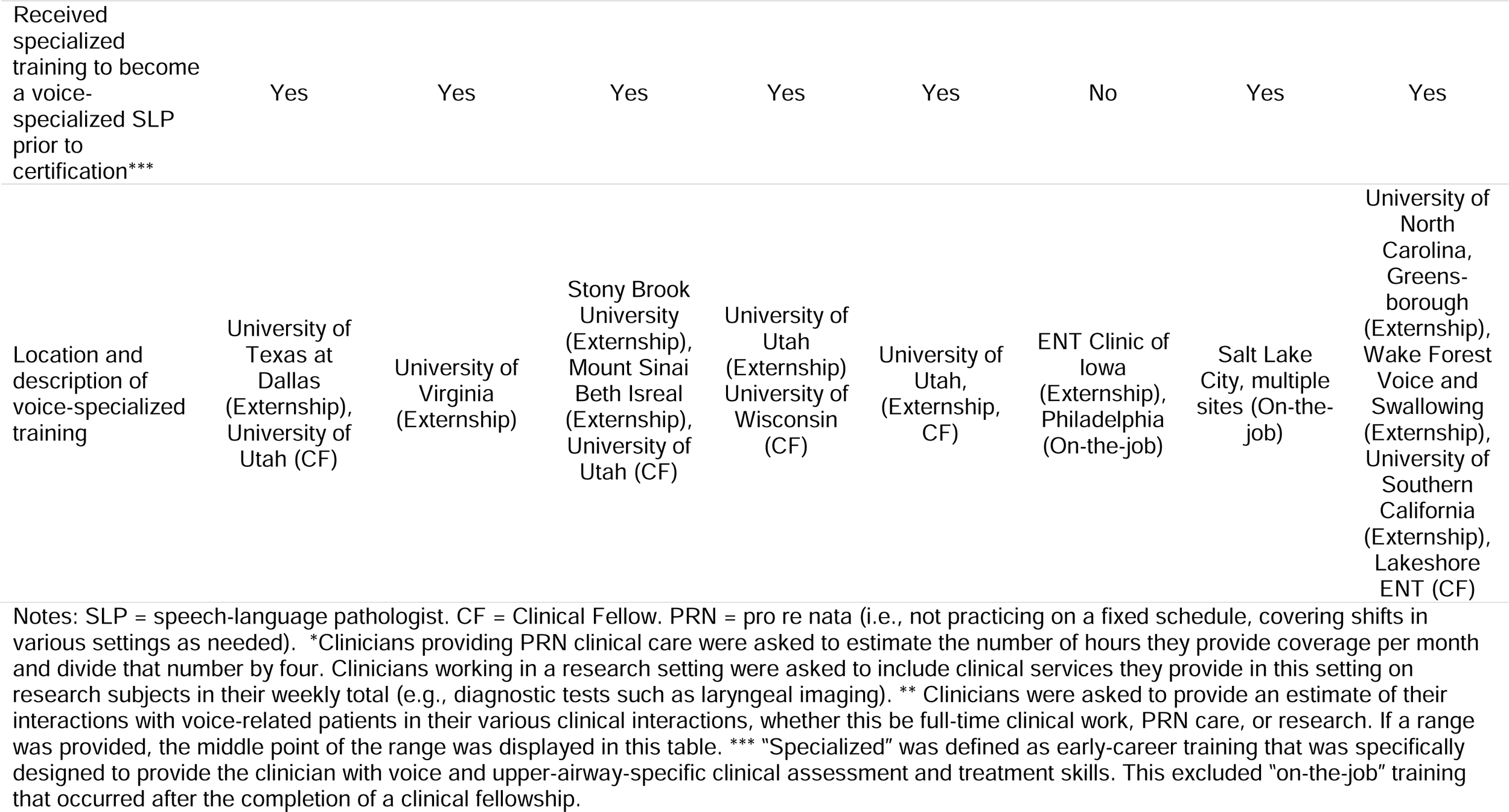
Clinician Demographics.

### Data Collection Procedures

Semi-structured interviews were used as the data collection method for this study. Each clinician participated in a semi-structured interview lasting approximately 30-45 minutes. This interview was conducted in person or via Zoom (Zoom Inc.), depending on the clinician’s preference. All interviews were recorded using a computer Zoom room, which allowed for the use of Zoom’s automatic transcription to transcribe interview utterances. The same interview protocol was used for all clinicians in each interview; these questions are provided below. The predetermined questions listed below were used to facilitate and guide the discussion; however, the author used prompts and follow-up questions as necessary to elicit thick descriptions of each clinician’s thoughts and descriptions of stimulability testing, stimulability level, and their clinical reasoning process surrounding using stimulability testing for patients with voice disorders. Clinicians were not provided with a definition of stimulability testing or stimulability level prior to the interview, although the author did discuss her viewpoints on the topic with the clinician during the interview as she deemed appropriate to help foster continued conversation.

The interview protocol, with the questions used to facilitate discussion during the interview are listed below. Numbered questions represent primary questions; alpha codes represent possible follow-up questions or prompts to probe for further discussion.

1. In your opinion, what is stimulability testing?

a. How do you define success during stimulability testing?
b. How do you apply the term in documentation?
c. If you had to give stimulability testing another name, what would that be?
2. In your opinion, what does it mean to be “stimulable”?
3. How do you typically assess for stimulability?

a. Tell me about your thinking process when you are planning stimulability testing.
b. How were you trained to think about this?
c. How has your clinical experience influenced how you think about this?
4. What are your goals during stimulability testing?
5. How do you know when you are “done” with stimulability testing?

### Data Analysis

#### Interview Transcription

The entirety of each interview was transcribed using Zoom’s automatic audio transcription capabilities and using a line-by-line coding for analysis. Volunteers went through each raw transcript (.txt) file and converted it into a word processing document containing a transcript of the interview text, including transcriptions of all audible speech, filler words (e.g., “um”, word repetitions) and nonverbals such as laughter. Each transcript was then double-checked for accuracy by the author, following which the data were transferred into Excel for analysis. Final transcripts contain line numbers for reference and a glossary of acronyms used throughout by speakers; all final transcripts are available in the supplemental materials at the following link: https://osf.io/f9pzk/overview

#### Interview Coding

The data for the interviews were coded following IPA methods outlined in Larkin et al. (2021), which is an inductive process involving multiple readings of the data, descriptive note-taking, and iterative development of themes considering the possible connection between emerging themes and subthemes. The first phase of coding utilized open coding to minimize coder bias and emphasize the clinician’s voice in the coding process. Open coding is the process of coding every section of the transcript with a label, or “code”, generated using inductive reasoning and created using words from the clinician’s transcript whenever possible (Strauss & Corbin, 1998). For example, a segment of an interview that read, “I mean, to some extent stimulability is like trial therapy” was coded as “line between stimulability testing and therapy.” A second round of coding was then completed using constant comparison of the codes to determine emerging or common abstract subthemes for individual clinicians. This round involved grouping codes with common elements into subthemes. This process was repeated to group common subthemes into themes. Triangulation (i.e., the comparison of emerging subthemes and themes across clinicians and across researchers [the author and D.Z., a qualitative researcher on the author’s dissertation committee]) was utilized across the first two clinicians to continue to refine and enhance the creation of both subthemes and themes. After the author felt confident that the emerging themes and subthemes from coding the first two clinicians were accurately capturing the contents of these two interviews, she proceeded to coding the remaining interviews.

## Results

Demographics of the eight clinicians are displayed in Table 1. Clinician names were replaced with randomly generated two-letter pseudo-initials to preserve clinician confidentiality. Interview transcripts are presented using ellipses “…” to denote pauses in speech and bracketed ellipses […] to denote where text was removed to improve the clarity of the message. Verbal fillers (e.g., “um”, “uh”, “like”), repeated words, and non-verbals such as laughter were the most frequently text in such instances, although other cuts were made for purposes of concision and general readability. The full interview transcripts, including all fillers and non-verbals, are available to any interested readers in the supplemental material.

Interpretative phenomenological analysis (IPA) of the eight interviews resulted in five themes: *(1) Perceiving and monitoring patient responses; (2) Developing and trusting clinical skills; (3) Decision-making strategies; (4) Drawing clinical conclusions;* and *(5) Fostering a purposeful therapeutic relationship.* These themes, their subthemes, and their relative distribution (representing the percentage of total codes that were categorized under that theme), are displayed in Table 2. *Developing and trusting clinical skills* was discussed the most frequently by clinicians and *Fostering a purposeful therapeutic relationship* was discussed the least often, although overall the distribution of all themes was relatively equal. The relative distribution of each theme by each clinician is displayed in Table 3.

**Table 2.** Themes and Subthemes Generated from Interpretive Phenomenological Analysis of Semi-structured Interviews, Including the Relative Distribution of Each Theme.

| Theme | Subthemes | Distribution |
| --- | --- | --- |
| Perceiving and monitoring patient responses | • Clinician-based versus patient-based responses<br>• Perceiving versus measuring patient responses | 19% |
| Developing and trusting clinical skills | • Developing clinical skills<br>• Trusting clinical skills | 25% |
| Decision-making strategies | • Determining the initial facilitator for stimulability testing<br>• Determining when to change facilitators during stimulability testing<br>• Determining when to stop stimulability testing | 21% |
| Drawing clinical conclusions | • Determining stimulability level<br>• Determining candidacy and prognosis for therapy | 23% |
| Fostering a purposeful therapeutic relationship | • Improving patient buy-in and self-efficacy<br>• Creating a safe therapeutic environment<br>• Tailoring the stimulability testing experience to patient needs | 13% |

**Table 3.** Distribution of Themes by Clinician.

| Theme | JL | RE | TT | WH | **EM** | **GA** | **HD** | **QD** |
| --- | --- | --- | --- | --- | --- | --- | --- | --- |
| Perceiving and monitoring patient responses | 9% | 25% | 14% | 19% | 17% | 25% | 25% | 17% |
| Developing and trusting clinical skills | 14% | 22% | 25% | 29% | 27% | 21% | 25% | 22% |
| Decision-making strategies | 21% | 15% | 25% | 29% | 28% | 24% | 11% | 18% |
| Drawing clinical conclusions | 48% | 32% | 17% | 12% | 17% | 15% | 21% | 32% |
| Fostering a purposeful therapeutic relationship | 9% | 5% | 19% | 12% | 11% | 16% | 18% | 11% |

2 Note: Bolded initials indicate the clinician is classified as late career.

### Perceiving and Monitoring Patient Responses

Both early- and late-career clinicians reported that perceiving and monitoring the patient’s response to stimulability testing was an essential element of their clinical reasoning and decision-making process. Within this theme, clinicians discussed measuring responses centered in the clinician’s perception as well as the patient’s perception of changes (Subtheme 1: Clinician-based versus patient-based responses) as well as differences in how patient response were measured (Subtheme 2: Perceiving versus measuring patient responses).

### Clinician-based versus patient-based responses

Clinicians reported monitoring a variety of patient behaviors during stimulability testing to inform their clinical reasoning. Most of these behaviors were centered in the clinician’s perception of the patient’s behaviors. For instance, all clinicians described monitoring the auditory-perceptual changes that occur in the patient’s voice quality during stimulability testing. Other clinician-based perceptual behaviors included the patient’s breath support, the presence or absence of perilaryngeal, neck, jaw, and upper body tension, the patient’s ability to follow directions, the patient’s ability to mimic clinician models, and the patient’s ability to control and coordinate laryngeal and speech musculature to produce the target sounds.

In addition, most clinicians also reported monitoring patient-based perception of changes during stimulability testing. Patient-based perception of vocal physiological effort, cognitive effort, and vocal pain were frequently mentioned. Assessing patient self-awareness or insight, which was generally described as the patient’s ability to hear or feel changes in their own voice, was noted as a strong priority for many clinicians. Clinicians described several strategies for assessing patient self-awareness, such as using nonverbal signs of frustration of confusion, directly asking the patient if they could feel or hear changes in their voice, or using the case history to determine how familiar the patient may be with their voice (e.g., “if we’re working with a singer, usually they’re going to have pretty high insight into the mechanism because of training”). Some clinicians additionally noted that they tried to ensure there was a match between their perception and the patient’s perception (“[I want to] see if the patient is in agreement with what my perception is”).

### Perceiving versus measuring patient responses

The strategies the clinicians used for evaluating patient responses during stimulability testing varied between clinicians. Most clinicians reported using an intuition-based approach, where they used an internal scale to track and measure change in response (i.e., *perceiving*). Some clinicians even reported that the degree of change they were seeking from patients on these scales varied depending on the patient presentation and the clinical goals the clinician had in mind:

I will use my clinical judgment to sort of decide like, is that enough improvement for me to feel like I’m accomplishing what I wanted to accomplish as a clinician […] it can depend on, on what we’re trying to target […] can I make it better? Did I make it better? Okay, great, we’re headed in the right direction.

A minority of clinicians, however, reported using external scales to gauge patient response to stimulability testing (i.e., *measuring*). One clinician reported a very structured stimulability process that utilized field-standardized auditory-perceptual evaluations of voice quality, acoustic measures of voice quality, and patient self-reports of voice quality, effort, and pain using clinic-standardized rating scales. This clinician reported doing “whatever I can do” to make their process of quantifying patient improvement “systematic” and “very deliberate”. Only one other clinician mentioned the use of objective measures of voice changes, such as acoustic or aerodynamic measures, during stimulability testing, and this clinician noted that they used these measures “only when I’m my best self”. The only other mentions of standardized measures of patient responses to stimulability testing was the use of standardized auditory-perceptual rating scales as an anchoring point for ratings of voice quality (two clinicians mentioned using the GRBAS scale; Hirano, 1981).

### Developing and Trusting Clinical Skills

The second theme, *Developing and trusting clinical skills,* speaks to the core technical skills that form the basis of stimulability testing. Clinicians discussed the process of developing (Subtheme 1) and then trusting (Subtheme 2) various clinical skills to perform effective stimulability testing within this theme.

### Developing Clinical Skills

Both early- and late-career clinicians discussed the development of their stimulability testing approach. Three distinct approaches towards performing stimulability testing emerged among the clinicians of this study. The first, termed a *framework-based* approach, was described as using an identifiable decision-making framework to help determine which facilitator and/or cueing method would likely be most beneficial throughout stimulability testing. The second, termed a *standardization-based* approach, was described as working sequentially through a standardized list of facilitators, specific to either a clinician or a workplace, usually in the same order, to determine which facilitator gives the patient the best benefit during stimulability testing. Finally, the *facilitator-based* approach was described as starting stimulability testing with the same facilitator with most patients, particularly those from the same diagnostic category (e.g., muscle tension dysphonia), with adjustments made to the use of that facilitator as needed.

Not all clinicians were aware of the contrasting approaches to stimulability testing, but many discussed the varying influences on how they approached stimulability testing. One clinician had trained at two facilities, one that used a standardization-based approach and one that used a framework-based approach, and discussed how both approaches had pros and cons. This clinician also expressed how, as a result, they utilized a blended approach: “[a framework-based approach] forces me to be more critical, and I prefer that. But then I do find like in situations where […] I maybe don’t have enough time […] I revert back to doing [a standardization-based approach]”. Another clinician also discussed how underlying treatment theories affected their decision to use a framework-based approach, stated that a standardization-based approach felt “against motor learning theory in my brain […] [because I’m] looking at true motor change [during stimulability testing]. Are they able to generate […] a voice beyond just a basic training gesture?” Thus, both training influences and clinician’s individual treatment theories behind stimulability testing can influence their selection of stimulability testing approach.

### Trusting Clinical Skills

All clinicians described how their chosen approach to stimulability testing increased their trust in their ability to perform stimulability testing adequately. For instance, a clinician who used the facilitator-based approach stated, “I feel like, like [my chosen facilitator] rarely doesn’t work, so I probably will just stick with that instead of trying other things”, while a clinician who favored a framework-based approach stated “the benefit to kind of picking, like two [facilitators] and going with it is it really forced me to be critical about ‘why am I choosing these stimulability tasks?” One clinician stated that projecting confidence in their clinical abilities was an integral element of their patients’ ability to improve during stimulability testing:

> if I’m not confident that they can, that their symptoms will respond […] or if they don’t think […] that I don’t think they can improve, I don’t think they’re likely to make significant gains.

### Decision-Making Strategies

Clinicians discussed the clinical reasoning and strategies used to make three primary decisions during stimulability testing: determining the initial facilitator for stimulability testing (Subtheme 1), determining when to change facilitators during stimulability testing (Subtheme 2), and determining when to stop stimulability testing (Subtheme 3).

### Determining the initial facilitator for stimulability testing

All clinicians discussed the reasoning process driving how to choose the initial facilitator when beginning stimulability testing. For clinicians who favored a standardization-based or facilitator-based approach, this decision was generally dictated by the respective approach.

Clinicians who described using a framework-based approach reported a more complex reasoning and subsequent decision-making process. Three different frameworks were reported by clinicians who utilized a framework-based approach; one clinician cited a published framework (Van Stan et al., 2015), another reported listening for whether it was “an airflow, a resonance, or a space issue”, and a third described trying to match patient presentation to underlying anatomy and physiology. These clinicians all described a reasoning process where an initial facilitator was selected based on the hypothesized underlying issue, using their respective framework to guide facilitator selection. For instance, the clinician who utilized the “space, resonance, airflow” framework described their approach as follows:

> if I’m seeing someone who’s really, really clenching their jaw, that might lead me to more of a space component, versus, you know, if they’re sitting in glottal fry the entire time, that might be more of an airflow thing […] if I have someone who I think is gonna need to work on airflow, I think there are certain tasks in terms of […] therapeutic approaches that I think are easier for patients to kind of notice the airflow. […] for instance, like all of our trills – you can’t do trills without airflow, right?

### Determining when to change facilitators during stimulability testing

Proponents of a standardization-based approach discussed that a primary benefit of this approach is it allowed them to trial multiple techniques to determine which gave the patient the “best bang for your buck”. In contrast, clinicians utilizing framework-based or facilitator-based approaches discussed a complex reasoning process regarding when to change facilitators when their initial choice of facilitator was not achieving the desired results. One clinician described a relatively structured approach to reasoning process, using a cut-off of approximately 60% accuracy to determine whether a facilitator is an appropriate fit for a patient before trialing another. Other clinicians discussed relying more heavily on patient feedback and monitoring for signs of frustration. Clinicians also described checking in with their own language and cueing techniques to determine whether they were the source of the disconnect. Finally, one clinician noted that if multiple facilitators were not achieving the desired results, they took this as an indicator as a need for more focused attention on a single facilitator (“after I do like 3 facilitators, and […] nothing’s really linked up for them, then I can usually know that we just need to spend some time.”)

### Determining when to stop stimulability testing

When queried, clinicians reported that stimulability testing took them anywhere from less than five minutes to 30 minutes to perform. Many reported time constraints as a contributing factor into how long they spent on stimulability testing. If given adequate time, clinicians reported the decision to terminate stimulability testing was based on when they felt they had accomplished their clinical goal(s). The goals of stimulability testing varied across clinicians and included a variety of clinician-directed (e.g., determine patient prognosis for behavioral therapy) and patient-directed (e.g., improve patient understanding of their voice condition) behaviors.

Some clinicians reported a “hierarchy” of goals that they hoped to achieve within stimulability testing, each taking progressively more time:

> Are they a candidate for therapy? I can tell that pretty quickly […] but then, if I have more time […] then you keep going and see if you can take their voice and stabilize it in conversational speech, um, to help them feel better that day […] That’s the goal is to get all of that done, but at minimum you want to know if they can make improvement in therapy.”

Similarly, another clinician discussed the decision to include or exclude stimulability testing from their evaluation procedures based on a similar hierarchy of therapeutic goals:

> [stimulability testing is] most important when you’re not sure if they’re gonna respond to therapy techniques […] like maybe someone with something we [speech-language pathologists] can’t fix, like a paralysis.

Thus, the decision to include, exclude, or halt stimulability testing involved a clinical reasoning process of determining clinical goals as well as which of these goals can be met within an allotted time frame.

### Drawing Clinical Conclusions

Clinicians reported using stimulability testing to determine stimulability level (i.e., the results of stimulability testing; the extent to which they described a patient as “stimulable”; Subtheme 1). Using stimulability level, clinicians could then draw conclusions about patient candidacy and prognosis for behavioral therapy (Subtheme 2).

### Determining Stimulability Level

Individual clinicians provided multiple different interpretations for how they determined stimulability level. Many clinicians described assessing at least two or three different factors concurrently when making a judgement of stimulability level, as illustrated in the following quote:

> a highly stimulable individual […] is able to mimic the targets that I give them through modeling […] and follow the directions […] I give them and make positive change to their vocal quality. […] the most highly stimulable are the ones that do that, and in the subsequent conversation it holds on, it stays, and it’s still present, at least for a short duration.

There was variability in whether clinicians discussed the weighting of various criteria in their determination of stimulability level. Some clinicians identified criteria that were weighed most heavily; for example, one clinician stated, “the main component would be their motivation”, and another stated “how quickly we’re able to get to a clear voice, which I think is more […] of what I put weight into as opposed to how many tasks they can be stimulable for.” Other clinicians, however, made no mention of weighting amongst the criteria that they listed as factors influencing stimulability level.

Stimulability level itself was also framed in several different ways, including as a binary, as a continuum, and both as a measure an individual’s overall potential change and as their potential for change for a specific facilitator. Generally, clinicians who used stimulability level as a binary did so when describing an individual, not their responses to a facilitator. Four of the eight clinicians generally used “stimulable/not stimulable” in this way. In contrast, two clinicians used the term “stimulable/not stimulable” for both facilitators and individuals. Conversely, one clinician used the term “stimulable” as a more continuous measure and described it as “a spectrum […] rather than yes or no, it’s highly stimulable to not at all stimulable”. Wording used to describe the scale used to judge stimulability level varied between clinicians, from generally vague descriptors such as “quite stimulable” to structured hierarchies, such as “high, medium, low” stimulability.

### Determining Candidacy and Prognosis for Therapy

While candidacy and prognosis are two separate phenomena, clinicians often talked about them jointly and they are thus discussed jointly. All clinicians discussed using stimulability testing to inform their judgment of either the patient’s candidacy for behavioral therapy or their prognosis for behavioral therapy once deemed a candidate. However, clinicians were split on whether they felt stimulability testing was truly able to determine patient candidacy or prognosis.

Two clinicians specified that patient candidacy for therapy is determined by whether the patient can achieve improvement in their voice symptoms with facilitative techniques during stimulability testing. One described this by stating, “can you improve their function?”, and the other “is their overall voice improving?”; if the answer was yes, the patient was deemed a candidate for voice therapy. Both clinicians also specified that this determination can be made extremely quickly, without patient input. However, these two clinicians also said that, if they have the time during stimulability testing, they prefer to spend it determining whether a patient has interest in what one clinician described as a “course of therapy” (which this clinician differentiated from “therapy techniques”), by assessing additional factors during stimulability testing such as the “self-efficacy portion […] all that metatherapy stuff”. These clinicians therefore drew a distinction between the ability for patients to change their voice (i.e., candidacy) and patients’ overall interest in engaging with the therapeutic process, although both factors could be determined using stimulability testing.

In contrast, three clinicians suggested a disconnect between stimulability testing and therapeutic candidacy and/or prognosis. Two of these clinicians discussed how the time frame of stimulability testing was not extended enough to truly determine a patient’s prognosis or candidacy for therapy. One late-career clinician expressed:

> …stimulability kind of lets me know my prognosis, but for me it […] doesn’t always give me a hard stop of like, ‘I’m not going to take on this patient’ because some people just need more time, more awareness building […] I still am a big believer in like at least two more sessions [of therapy beyond stimulability testing], because I think, to put that much pressure on a patient that it’s like ‘you have 10 minutes to perfectly find your voice […] that feels against motor learning theory to me.

Similarly, another clinician discussed the need for a “full hour therapy session” to truly determine a patient’s prognosis. One clinician also noted that sometimes stimulability testing gives them an impression of patient prognosis that is contrary to what they actually experience in therapy: “you’re like, oh, yeah, patient’s a great candidate based on, you know, 10 minutes of stimulability testing in their eval, and then you get to their first therapy session and you’re like, oh […] this is gonna be hard”. These three clinicians therefore described the difficulty in using an abbreviated evaluation such as stimulability testing to make strong conclusions regarding candidacy or prognosis.

### Fostering a Purposeful Therapeutic Relationship

All clinicians described purposefully connecting with their patients in goal-driven ways to improve the patients’ therapeutic outcomes both during and following stimulability testing. Clinicians described forming therapeutic relationship with patients during the stimulability testing process to improve both patient buy-in and self-efficacy for voice therapy (Subtheme 1), as well as to create a sense of safety in the therapeutic environment (Subtheme 2) and tailor the stimulability experience to each patient’s individual needs (Subtheme 3).

### Improving Patient Buy-In and Self-Efficacy

Many clinicians discussed how stimulability testing acted as “teaser” for voice therapy that helped both the clinician and the patient to determine and refine their interest in pursuing a course of behavioral therapy. By using stimulability testing in this way, clinicians described shifting the decision regarding whether to engage in therapy to the patients (“…look! We can make changes with your voice. This is what therapy would look like. Is this something you want to do?”). Some clinicians described helping patients to achieve self-efficacy and even self-sufficiency with voice changes during stimulability testing as their primary goal of performing stimulability testing:

> But to really get them to buy in, that’s the other thing to me. […] are we just determining candidacy for therapy? […] why are we doing this? If we’re just trying to determine therapy candidacy, then to me […] that could be done very quickly. If we want buy in, we want them to get better soon, now, we’re going to stimulate that improvement […] and see if they can actually sustain it.

### Creating a Safe Therapeutic Environment

Clinicians universally demonstrated a desire to provide patients with a stimulability testing experience that encourages, rather than discourages, participation in voice therapy. They did so by discussing their strategies for minimizing or reducing patient overwhelm, frustration, or confusion during stimulability testing. Many clinicians endorsed having a “script” that they used at the beginning of stimulability testing to prepare patients for the stimulability testing process. One clinician described checking in with patients regarding their readiness to achieve normal voicing during stimulability testing within this script (“There’s a really good chance we can get your voice back today […] Do you have any objections to that?”) to provide space for patients who may have barriers to achieving or thinking they can achieve such a goal during the voice evaluation. Clinicians also endorsed frequently evaluating patients throughout stimulability testing, both verbally and by assessing non-verbal cues, to gauge the degree of patient engagement. Many clinicians described the patient experience as taking precedence over clinician perceptions (“I’m not gonna keep pushing them, even though I may think they’re capable of more improvement”) due to the importance of keeping a patient engaged in the therapeutic process (“they get frustrated like, you’re not really helping them that much, and then they’re turned off by the whole idea of therapy”).

### Tailoring the Stimulability Testing Experience to Patient Needs

Clinicians also discussed tailoring their stimulability testing protocols to the needs of individual patients. Frequently, this took the form of integrating the individual medical, social, and occupational needs of a patient into their plan of care and adjusting stimulability testing accordingly. One clinician summarized an example of how they might tailor stimulability testing to suit the needs of a patient as follows:

> …it may even be that it is a really complex […] medical condition, and it really does affect them, and they do want to change, but they just might be so tired from all the other things they have going on […] It’s just kind of figuring out […] what is one thing I can do to try and get you a little better […] Instead of doing a whole […] thing of resonant voice, or even doing just the different pitch glides and hums […] just can you hum for me?

Additionally, some clinicians also discussed trialing different techniques simply because “what is going to click for someone […] may not click for the next person”. For example, one clinician talked extensively about using their framework-based approach to guide them through trialing various ways of cueing the patient to find an approach that worked: “Okay, somatosensory is not really resonating with them, maybe we move to like pain monitoring.”

## Discussion

The current study is the first to examine the clinical reasoning process during stimulability testing completed by voice-specialized speech-language pathologists. Three broad frameworks for clinical reasoning exist in the literature: analytical frameworks, which involve a conscious, deliberate process of analyzing incoming information and comparing it to patterns in a mental representation of a diagnostic category or other clinically useful standard; intuitive frameworks, which rely primarily on subconscious information-gathering that occur during patient-provider interactions; and hybrid, or combination, frameworks, which integrate elements from both frameworks. Qualitative analyses using an interpretative phenomenological analysis methodology yielded five themes, which provided excellent insight into the clinical reasoning process occurring during stimulability testing. In addition, these themes also reflect the enormous complexity of stimulability testing as a clinical skill for voice-specialized speech-language pathologists.

The five themes suggest that clinicians in the current study utilize a combination framework of clinical reasoning. The themes included both largely analytical processes (*Decision-making strategies*), largely intuitive processes (*Fostering a purposeful therapeutic relationship*), and processes that could be either analytical or intuitive, depending on the clinician or situation (*Perceiving and monitoring patient responses*). This finding was unsurprising in that it aligned with both the contextualized clinical reasoning framework, a framework specific to speech-language pathology (Pillay & Pillay, 2021) and the overarching framework for evidence-based clinical practice provided by ASHA. Given the lack of systematic research on stimulability testing, it was encouraging that clinicians were using an analytical, evidence-based approach (insofar as it was possible) rather than relying solely on intuition-based practices while performing stimulability testing.

Certain clinicians did appear to rely more heavily on an analytical or intuitive approach to stimulability testing. Of all the interviewed clinicians, HD used the most structured, quantitative approach to evaluate patients’ symptoms during stimulability testing, as well as a heavily complaint-focused style of performing stimulability testing, both of which suggest a clinical reasoning approach that is weighted more heavily towards analytical reasoning. In contrast, clinician TT, as a relatively new clinician, spoke frequently about using stimulability testing as an information-gathering tool as they expanded their own clinical toolbox, as well as a chance to connect with the patient and determine their goals for therapy, suggesting a more intuitive approach. This contrasts somewhat with previous literature regarding clinical reasoning development, which suggests that intuitive reasoning is generally a skill favored by expert clinicians due to its heavy reliance on experience-based pattern recognition (Banning, 2008). Despite this, as seen in Table 3, all clinicians discussed all five themes, indicating that every clinician engaged in a combination approach to at least some extent.

Each theme also provided unique insights into the clinical reasoning processes occurring during stimulability testing. Within the *Perceiving and monitoring patient responses* theme, clinicians discussed evaluating and integrating information from multiple sources. Both the *Developing and trusting clinical skills* and the *Decision-making strategies* themes spoke to the extremely complex nature of stimulability testing, outlining multiple diverse approaches to the assessment procedure across clinicians even within a relatively limited geographic sampling location (Salt Lake City, Utah). The lack of standardization in stimulability testing approaches resulted in a heavy reliance on a clinician’s training and intuition for the many clinical decisions required throughout stimulability testing. Regardless of the approach to stimulability testing, the *Fostering a purposeful therapeutic relationship* theme demonstrated the commitment of all clinicians to centering stimulability testing around the patient’s needs and wellbeing.

Interestingly, the results of the *Drawing clinical conclusions* theme suggested disagreement among the clinicians on two critical factors: the description of stimulability level (i.e., the results of stimulability testing; the extent to which a patient was considered “stimulable”), and the ability of stimulability testing to provide meaningful information regarding patient prognosis and candidacy for behavioral therapy. Clinicians described a variety of factors that influenced their judgement of stimulability level and varied in whether they discussed differential weighting of these factors into their final determination of stimulability level.

Additionally, clinicians variably used binary, categorical, and continuous measurement scales to describe stimulability level, and used stimulability level to describe both person-specific (e.g., “they are stimulable for therapy”) and behavior-specific (e.g., “stimulable for resonant voicing”) outcomes. As individuals, clinicians explained the reasoning behind their formulation of stimulability level adequately; however, as a group, there was minimal consensus on what constituted stimulability level. Perhaps because of this, clinicians did not draw the same conclusions regarding the relationship between stimulability testing and patient prognosis for behavioral therapy. In fact, nearly half of the clinicians (3/8) suggested that stimulability testing was not sufficient to gauge patient prognosis for behavioral therapy. This represents a strong contrast with previous work, where 100% of surveyed clinicians reported that stimulability testing predicted overall therapy outcome at least sometimes (Toles & Young, 2023).

These findings suggest that the complex, unstandardized methods of stimulability testing represent a barrier to consensus in clinical decision-making amongst practicing voice-specialized speech-language pathologists. While the clinical reasoning *process* described within this study was well-formed and well-described for many clinicians, the resulting clinical *decisions* lacked uniformity and confidence. This likely reflects the lack of evidence-based knowledge regarding the outcomes of stimulability testing in the current literature. While stimulability level has been described as a prognostic indicator in the speech sound disorders literature (e.g., Powell & Miccio, 1996), there is extremely limited literature linking stimulability level to prognosis in patients with voice disorders (Dejonckere & Lebacq, 2001; Shelly et al., 2023). Additionally, stimulability level is variably described in the literature; in a scoping review, 12% of articles described stimulability level as an individual-specific measure (e.g., “stimulable for therapy”), 78% described it as a behavior-specific measure (e.g., “stimulable for resonant voicing”), and 9% described it as a both an individual- and behavior-specific measure (E. D. Young, 2025). This same review found that stimulability level was described using a binary measurement scale in 78% of articles (e.g., “stimulable” vs “not stimulable”), a categorical scale in 15% of articles (e.g., “excellent, good, or poor stimulability”), and using multiple measurement scale types in 7% of articles. Thus, the established literature provides limited and conflicting guidance for practicing clinicians regarding clinical decisions following stimulability testing.

The lack of established guidelines for stimulability testing resulted in a heavy reliance on drawing intuition-based conclusions following stimulability testing. While intuition-based clinical decision-making undoubtedly has its place in stimulability testing, an overreliance on intuition for drawing clinical conclusions suggests that more research is needed to elucidate the relationship between stimulability testing, stimulability level, and prognosis and candidacy for behavioral therapy. Several authors have recently advocated for standardizing stimulability testing procedures in response to the variability that exists in clinical practice (Petty et al., 2023; Toles & Young, 2023; Weston et al., 2023). An additional option for addressing these concerns is to create a standardized framework for describing both stimulability testing and reporting stimulability level in research and clinical practice. Both options underline the need for additional research upon which practicing speech-language pathologists can draw to inform their clinical reasoning when performing stimulability testing. Without this work, practicing clinicians will continue to rely solely on intuition-based reasoning, which will likely continue to result in poor consensus in their subsequent clinical decisions between clinicians.

An additional benefit of increased systematic research into stimulability testing and stimulability level is an increased ability to effectively educate new clinicians about how to perform the practice. It was notable that the *Developing and trusting clinical skills* theme was the most discussed of all themes, underling the tremendous clinical, cognitive, and emotional work that goes into learning how to effectively implement these skills. Clinicians within the current study described learning how to perform stimulability testing through direct supervision and teaching, observation of other clinicians, and repetition and practice. The creation of standardized frameworks of stimulability testing and stimulability level would allow movement away from a mentorship-style model, making the skill more accessible to clinicians who do not have access to one-on-one mentorship with a voice-specialized mentor. Standardization would also help to make stimulability testing less of a “black box” for new clinicians, providing them with increased support in the clinical decision-making process during the early stages of their career. Such scaffolding could provide a much-needed boost to early-career clinicians to improve not only their confidence but competence in stimulability testing.

The current study does have some limitations. First and foremost, the sample of clinicians for was limited both in terms of geographic region and in clinical practice sites. While purposefully selected to be from a variety of training levels and with the intent of achieving as wide a range of training backgrounds as possible, the clinicians within the current study are all from an extremely tight-knit group of voice-specialized speech-language pathologists in the Salt Lake area. Specialization in voice disorders is relatively uncommon amongst speech-language pathologists, and as such it is very difficult to find a sample of voice-specialized speech language pathologists in the same geographical location that do not know each other, work together, or have trained one another. Thus, the present single-site data collection strategy made it nearly impossible to control for the influence of training background or workplace.

Collecting data from clinicians from multiple different sites would have yielded a much broader range of perspectives on stimulability testing, and potentially a very different pattern of results. Indeed, this is a potentially fruitful avenue for future research. Second, as stimulability testing is a process that can depend greatly on patient presentation, some clinicians had difficulty describing their stimulability testing approach in an open-ended interview format. Either presenting clinicians with standardized case studies or interviewing clinicians after performing stimulability testing could be potential methods of future study for further evaluating clinical reasoning in stimulability testing.

## Conclusion

Stimulability testing is a valuable but nuanced skill that involves complex clinical reasoning processes. The current study represents the first examination of how speech-language pathologists use stimulability testing to inform clinical decisions such as judgements of stimulability level and the determination of patient candidacy and prognosis for behavioral therapy. Clinicians relied heavily on intuitive reasoning for drawing these clinical conclusions, likely because of a lack of systematic research tying stimulability testing to outcomes such as patient candidacy and prognosis. More research is needed to continue to explore how clinicians

## Data Availability

The datasets generated during an analyzed during the current study are available in the open science framework repository, https://osf.io/f9pzk/overview

## Acknowledgements

Special thanks to Sarah Hargus Ferguson, Julie Barkmeier-Kraemer, Nelson Roy, Adrianna Shembel, and Donna Ziegenfuss for their valuable feedback on original versions of this manuscript. Particular thanks go to Donna Ziegenfuss for her guidance and help with the analysis throughout this project. A huge thank you to Alexis Bitner, Blake Bradley, Angie Gomez, Hartley Hartley, Anna Humphries, Camrynn Nalwalker, and Celia Young for their hard work and countless hours spent revising the original transcripts for this project. The author’s positionality statement is available at the following link: https://osf.io/f9pzk/overview

## Notes

### Competing Interest Statement

The authors have declared no competing interest.

### Funding Statement

This project was funded through the University of Utah CSD Department Seed Grant for Innovative Research.

### Author Declarations

IRB of the University of Utah gave ethical approval for this work

